# Non-invasive prehabilitation before neurosurgery modifies the topography of brain language networks without compromising function

**DOI:** 10.64898/2026.04.13.26350473

**Authors:** Nuria Brault-Boixader, Alba Roca-Ventura, Selma Delgado-Gallén, Edgar Buloz-Osorio, Leonardo Boccuni, Carlos Laredo, Emma Muñoz-Moreno, Núria Bargalló, David Bartrés-Faz, Álvaro Pascual-Leone, José M. Tormos-Muñoz, Ruben Perellón-Alfonso, Kilian Abellaneda-Pérez, Prehabilita Working Group

## Abstract

Patients with brain tumors involving language-critical regions face surgical limitations when balancing resection with preservation of function. Non-invasive neuromodulation-induced prehabilitation (NIP) aims to guide preoperative neuroplastic reorganization, potentially facilitating larger resections while preserving function. We investigated whether NIP selectively modulates the targeted language network compared with control networks, and whether such modulation is behaviorally safe. We enrolled 26 patients (mean age = 55.9 ± 11.8 years) from the Prehabilita project (Clinical Trial: NCT05844605) with operable brain tumors affecting language or motor regions. Eleven received language-targeted NIP, combining transcranial magnetic stimulation and/or transcranial direct current stimulation with intensive language training. Fourteen patients with NIP targeting non-language networks, primarily motor networks, served controls. Assessments included task-based functional magnetic resonance imaging (tb-fMRI) and a neuropsychological battery assessing language and cognitive domains before and after prehabilitation. Results indicated a group-specific NIP effect on the language network. In the language-targeted group, tb-fMRI revealed reduced overlap between a region of interest centered on the stimulation target and fMRI-derived language activation maps, whereas no comparable changes were observed in controls. No significant modulation effects were detected in the motor network in either group. These findings indicate that NIP can selectively reorganize the language network, with modulation patterns differing in sensorimotor networks. Importantly, language network modulation occurred while preserving language and cognitive performance. These results support NIP targeting higher-order functions such as language as a safe preoperative strategy that may reduce functional constraints on surgery and enable larger and safer resections in patients with tumors involving language-critical regions.

## 1. Introduction

Cancers of the central nervous system (CNS) represent a complex group of neoplasms originating from diverse cell types within the brain, spinal cord, and surrounding tissues (Salari et al., 2023; PDQ Adult Treatment Editorial Board, 2024). These tumors are a leading cause of morbidity, severe disability, and mortality, imposing a significant burden on healthcare systems (Deorah et al., 2006; Fan et al., 2022). According to the most recent reports (Bray et al., 2024), in 2022 there were 321,476 newly diagnosed cases of CNS cancers, causing 248,305 deaths worldwide. Despite advances in the fields of neurosurgery, radiotherapy, and chemotherapy, the overall 5-year survival rate for malignant brain tumors is estimated to be 35.7% (Ostrom et al., 2023). In the context of brain tumor surgery, patient survival and quality of life depend not only on the extent of tumor resection, but also on preserving healthy brain tissue and neurological function (Brown et al., 2016; Roh & Kim, 2023). However, the balance between tumor removal and the preservation of functional tissue is often challenging, particularly when tumors are located within or invade brain regions holding critical functions, such as those involved in language (Shah et al., 2025).

Language is a fundamental human cognitive function, subserved by a large-scale neural network characterized by dynamic interactions among distributed brain regions, mainly including the frontal and temporoparietal cortices (Hickok & Poeppel, 2007; Nieberlein et al., 2023). The language system in the brain is highly plastic, as evidenced by its capacity for adaptive reorganization after injuries to mitigate language-related impairments, such as those caused by brain tumors (Fisicaro et al., 2016) or following a stroke (Geranmayeh et al., 2014; Tilton-Bolowsky et al., 2024). Such reorganization includes recruitment of distinct perilesional regions, the activation of homologous areas in the contralateral hemisphere, as well as the engagement of domain-general regions in both hemispheres (i.e., the prefrontal cortex; Cirillo et al., 2019; Fedorenko et al., 2024; Geranmayeh et al., 2014; Martin et al., 2022; Nieberlein et al., 2023).

In this context, non-invasive brain stimulation (NIBS) techniques, which can be used to induce and guide brain plasticity processes (Jannati et al., 2023; Kirton et al., 2021), can be powerful allies. Previous research has consistently shown that NIBS protocols can modulate functional activity and connectivity across key brain networks in healthy individuals (e.g., Abellaneda-Pérez et al., 2019; Halko et al., 2014; Wang et al., 2014), including the language network (e.g., Meinzer et al., 2013). Moreover, NIBS has also been shown to influence network patterns in distinct neurologic and psychiatric conditions (e.g., Liston et al., 2014), including those within the language network (Meinzer et al., 2015). Building on this evidence, NIBS is emerging as a promising interventional approach for pre-surgical conditioning in brain tumor patients, seeking to reconfigure critical brain networks before surgery to enable safer and more extensive resections (e.g., Barcia, Sanz, González-Hidalgo, et al., 2012; Rivera-Rivera et al., 2017; Dadario et al., 2022; Lang et al., 2020; Engelhardt, 2023). These approaches are known as neuromodulation-induced prehabilitation (NIP) and are based on combining non-invasive brain stimulation (NIBS) with intensive, function-specific training. A key feature of NIP is the pairing of inhibitory-like neuromodulation applied to peritumoral regions with targeted behavioral training delivered during or immediately after brain stimulation, with the aim of transiently suppressing local activity while exploiting windows of heightened neuroplasticity to facilitate compensatory network recruitment and adaptive functional reorganization (Bolognini, 2009). Ultimately, these approaches aim to enable more extensive tumor resection (potentially reducing recurrence risk) without increasing the likelihood of postoperative functional deficits (Boccuni et al., 2023). Recently, we applied a non-invasive NIP protocol, combining NIBS with intensive training, in a case series of ten patients, initially showing that these protocols are feasible, safe and well tolerated (Boccuni et al., 2024a). Moreover, in an fMRI-based case report, we provided a detailed characterization of the potential neural mechanisms underlying this NIP process within the language network (Boccuni et al., 2024b). Altogether, these findings highlight the potential for language-network plasticity prior neurosurgery, revealing substantial capacity for functional modulation.

Nonetheless, NIP research remains limited and largely focused on clinical outcomes (Barcia et al., 2012; Boccuni et al., 2024a; Boccuni et al., 2024b; Dadario et al., 2022; Ekert et al., 2024; Lang et al., 2020; Serrano-Castro et al., 2020), leaving the neural mechanisms underlying these changes largely unexplored. In this vein, systematic characterization of NIP-associated network-level topological changes and their relationship to behavioral outcomes remains largely lacking, particularly in cognitive and language-related NIP protocols (e.g., Dadario et al., 2022; Lang et al., 2020; Ekert et al., 2024). This gap is of central importance, as tumors located near language regions are frequently considered inoperable due to the risk of intervention-related language impairment (Collée et al., 2022; Sanai & Berger, 2008). Indeed, postoperative communication deficits are among the most disabling outcomes of brain surgery, with major consequences for independence and quality of life and they can persist even when function-preserving approaches such as awake mapping are used (Collée et al., 2022; Vogrincic et al., 2025). Furthermore, language preservation is a cross-cutting neurosurgical problem extending beyond neuro-oncology (e.g., epilepsy surgery), reinforcing the broader relevance of understanding language-network plasticity and its modulation for distinct translational clinical applications (Zhu et al., 2025). Accordingly, the present study focused on comprehensively characterizing these network-level effects in a larger, more homogeneous cohort and comparing them with a non-language NIP control group.

Specifically, this study aims to: (1) examine whether NIP protocols targeting the language network (as defined by task-based fMRI activation maps), elicit brain network changes that are specific to the targeted system (i.e., language network) and are dissociable from modifications in non-targeted brain networks (i.e. motor network); (2) assess whether these language-network changes are associated with measurable behavioral modifications in language performance during NIP intervention; and (3) determine whether these neural and behavioral NIP-related modulations occurs with or without entailing deleterious effects on other cognitive processes (i.e., potential crowding effects). We hypothesize that non-invasive NIP will induce specific topographical reorganization within the targeted language network, reflected in measurable pre vs. post intervention shifts in tb-fMRI activation patterns. Considering the distributed and multimodal architecture of the language network, we hypothesize that induced neural changes will remain behaviorally neutral, with no measurable detrimental effects on language performance or other cognitive domains (e.g., visuospatial constructional and perceptual abilities).

## 2. Materials and methods

This study is part of the Prehabilita project, conducted at Institut Guttmann (Guttmann Barcelona, Brain Health and Neurorehabilitation, Barcelona, Spain). A detailed description of the prehabilitation protocol employed in this study can be found in Boccuni et al. (2023).

### 2.1 Participants

A total of 30 patients were enrolled in the prehabilitation program. This cohort included participants recruited within the ongoing clinical trial (ClinicalTrials.gov identifier: NCT05844605) as well as individuals enrolled during a preceding pilot phase of the project. These patients were awaiting neurosurgery for a brain tumor affecting language or other non-language functions. Patients were referred from various Neurosurgery Units at different Hospitals across Catalonia, Spain. Exclusion criteria included contraindications for NIBS protocols, unstable medical conditions, a history of substance abuse, and severe musculoskeletal or cognitive disorders that could impact the intervention. Of the initial 30 participants, 4 were excluded from the current study: one due to fMRI acquisition issues; two because their tumors had non-language or non-motor functional involvement; and one because stimulation was delivered altogether to two functionally distinct targets (i.e., motor and language areas). The final sample comprised 26 patients (10 women; mean age = 55.88 ± 11.81 years), 12 of whom had high-grade gliomas (Table 1). Moreover, one participant who received only multifocal tDCS was excluded from one specific fMRI analysis (“2.6.1 Local effects of NIP at the stimulation target”) because an individualized ROI could not be defined. Additionally, one participant in the control group was excluded from the same analysis and another (“2.6.2 Effects of NIP on targeted network size”) due to missing post-intervention neuroimaging data on hand function.

**Table 1.** Characteristics of study participants in each group (age, sex, patient symptoms, tumor diagnosis, tumor side, NIBS protocol). L/R: Left or Right; NIBS: non-invasive brain stimulation. RLL: Right lower limb; LRL: Left lower limb.

| Language group (n = 11) |  |  |  |  |
| --- | --- | --- | --- | --- |
| ID | PATIENT SYMPTOMS | TUMOR DIAGNOSIS | MAIN TUMOR LOCATION (LOBE, SIDE) | NIBS PROTOCOL |
| 1 | Comitial crisis | Astrocytoma (WHO grade IV) | Frontal, L | 1hz rTMS + multifocal tDCS |
| 2 | Mild speech disturbance | Cavernoma | Parietal, L | 1hz rTMS |
| 3 | Mild speech disturbance (nomination, repetition) / Mild memory impairment (short term, working memory) | Cavernoma | Temporal, L | 1hz rTMS |
| 4 | Mild speech disturbance / Mild memory impairment / Partial comitial crisis (olfactory crisis) | Astrocytoma (WHO grade III) | Frontal & Temporal, R | 1hz rTMS + multifocal tDCS |
| 5 | Mild speech disturbance | Glioblastoma (WHO grade IV) | Frontal, L | 1hz rTMS |
| 6 | Mild speech disturbance | Glioma | Frontal, L | 1hz rTMS + multifocal tDCS |
| 7 | Partial comitial crisis (olfactory crisis) | Ganglioglioma (WHO grade I) | Frontal, R | 1hz rTMS + multifocal tDCS |
| 8 | Partial focal crisis in face and neck / Paraphasia's | Glioblastoma (WHO grade IV) | Frontal, L | 1hz rTMS + multifocal tDCS |
| 9 | Tonic-clonic seizures, paraphasia's | Glioblastoma (WHO grade IV) | Frontal, L | 1hz rTMS + multifocal tDCS |
| 10 | Aphasia, paraphasia, mild cognitive impairment | Glioblastoma (WHO grade IV) | Temporal, L | 1hz rTMS |
| 11 | Paraphasia, mild cognitive impairment | Oligodendroglioma (WHO grade II) | Frontal, L | 1hz rTMS + multifocal tDCS |

| Non-language group (n = 15) |  |  |  |  |
| --- | --- | --- | --- | --- |
| ID | PATIENT SYMPTOMS | TUMOR DIAGNOSIS | TUMOR LOCATION (LOBE, SIDE) | NIBS PROTOCOL |
| 12 | Moderate-severe right-hand motor and sensory impairment | Meningioma (WHO grade I) | Parietal, L | 1hz rTMS |
| 13 | Transitory left facial supranuclear paralysis / Involuntary movements / Speech disturbance | Glioblastoma (WHO grade IV) | Frontal, R | 1hz rTMS |
| 14 | Dizziness / Presyncope | Astrocytoma (WHO grade II) | Frontal, R | multifocal tDCS |
| 15 | Comitial crisis | Astrocytoma (WHO grade II) | Frontal, R | 1hz rTMS |
| 16 | Left hand sensory impairment (paraesthesia) | Meningioma (WHO grade II) | Parietal, R | 1hz rTMS |
| 17 | Comitial crisis | Oligodendroglioma (WHO grade II) | Frontal, R | 1hz rTMS |
| 18 | Left hand sensory and motor impairment | Glioblastoma (WHO grade IV) | Frontal, R | 1hz rTMS |
| 19 | Left hand sensory and motor impairment (paraesthesia, clumsiness and reduced grip) | Glioblastoma (WHO grade IV) | Frontal, R | 1hz rTMS + multifocal tDCS |
| 20 | Moderate left lower limb motor impairment (apraxia) | MT: Non-Small Cell Carcinoma | Frontal, L | 1hz rTMS + multifocal tDCS |
| 21 | Mild right hemiparesis / RLL Hypoesthesia and Dysesthesia / Mild motor impairment in RLL / attention deficits | Astrocytoma (WHO grade II) | Frontal & Parietal, L | 1hz rTMS |
| 22 | Tonic-clonic seizures | Glioblastoma (WHO grade IV) | Frontal, L | 1hz rTMS + multifocal tDCS |
| 23 | Left frontoparietal haemorrhagic stroke, mild right hemiparesis, mild cognitive impairment | Oligodendroglioma (WHO grade III) | Frontal & Parietal, L | 1hz rTMS |
| 24 | LRL paresthesia / Focal crisis | Glioblastoma (WHO grade IV) | Frontal, L | 1hz rTMS |
| 25 | RLL motor impairment | Meningioma (WHO grade I) | Frontal, L | 1hz rTMS |
| 26 | Left arm paresthesia, hand fine motor impairment | Gliosarcoma (WHO grade IV) | Frontal, R | 1hz rTMS |

Patients underwent comprehensive clinical, cognitive, and motor assessments, as well as all the prehabilitation intervention sessions, at Institut Guttmann in Barcelona, Spain. Neuroimaging assessments were performed at the IDIBAPS Magnetic Resonance Imaging Core Facility at Hospital Clinic de Barcelona, Barcelona, Spain. Recruitment spanned from July 2021 to December 2024, with evaluations conducted at two different time-points: before and after NIP. See Fig. 1 for an overview of the study design and timeline. Informed consent was obtained from all participants. All procedures from the present investigation were performed in accordance with the declaration of Helsinki. The study was approved by the Research Ethical Committee of Fundació Unió Catalana d’Hospitals (approval number: CEI 21/65, version 1, 13/07/2021).

**Figure 1.**
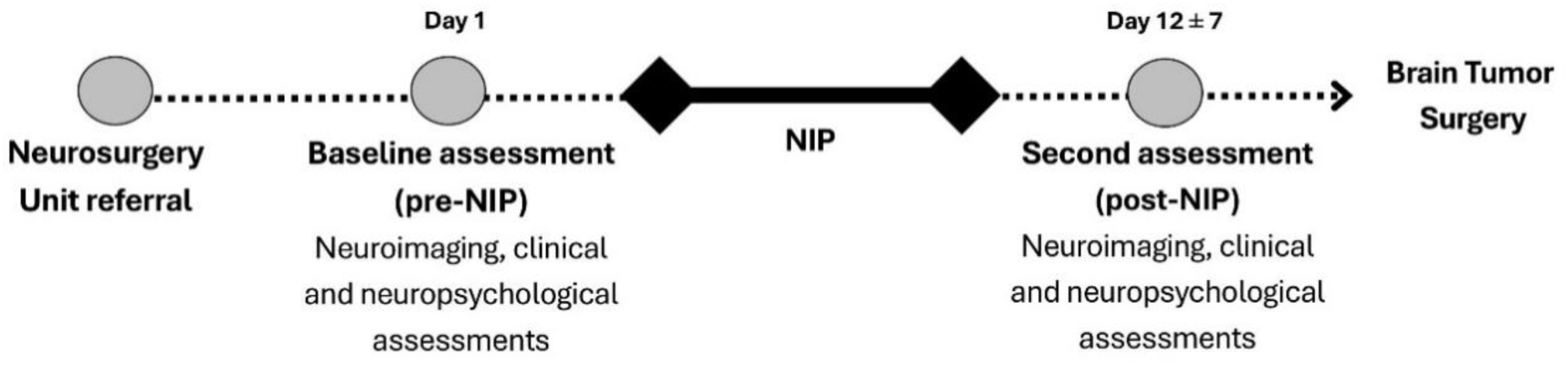
Study design and timeline, outlining the neuroimaging, clinical and neuropsychological assessments conducted pre- and post-NIP. The prehabilitation program lasted 12 ± 7 days, and surgery was performed 3 ± 2 days after completion.

### 2.2 Clinical, neuropsychological and motor assessments

Comprehensive clinical and neuropsychological assessments were conducted at each time point (pre-NIP and post-NIP) for all patients (see Fig. 1). Evaluations of language and cognitive functions were performed by licensed clinical neuropsychologists, while motor function examinations were performed by physiotherapists specialized in neurorehabilitation. For further details on the study assessments, the reader is referred to Boccuni et al. (2023). For this investigation, we focused specifically on data related to the NIP intervention to evaluate the immediate neural and behavioral effects of the prehabilitation process. Postoperative outcomes were beyond the scope of this analysis, which was designed to isolate the direct effects of prehabilitation.

#### 2.2.1. Language and neuropsychological functioning

The neuropsychological assessment used was the Barcelona-Revised Test (TB-R; Peña-Casanova, 2005). This battery of tests includes spontaneous language (conversation and narration, thematic narration, and picture description); fluency and informative content (qualitatively assessed based on the information obtained in the spontaneous language subtest); verbal word repetition; verbal naming (i.e., response by naming); verbal comprehension; reading comprehension (using sentences and texts); automatic language (using both forward and reverse sequential series); verbal repetition (using syllables, logatomes, and sentences); naming (using visuo-verbal naming and naming completion); and verbal comprehension (complex verbal material).

Moreover, to examine whether the functional impact of the prehabilitation-induced brain reorganizations extended beyond the targeted language domain to other cognitive functions (i.e., non-language-related processes), this investigation included neuropsychological assessments of visuospatial constructional and perceptual abilities. These were evaluated using the Block Design subtest of the WAIS-III (Wechsler, 2008) and the visuospatial attention subtest of the TB-R test.

### 2.3 Neuroimaging acquisition and analysis

#### 2.3.1 MRI acquisition

MRI data was acquired as part of the general NIP protocol (Fig. 1) using a 3-T Siemens scanner (MAGNETOM Prisma) equipped with a 64-channel head coil, except when the patient doesn’t fit comfortably; in those cases, we use the more spacious 20-channel coil. As detailed in the protocol description (see Section 2.3; Boccuni et al., 2023), comprehensive MRI acquisitions, including structural, diffusion, task-based, and resting-state sequences, were performed before and after NIP. For the purposes of the present study, we focused exclusively on structural and task-based fMRI data.

For structural imaging data, a high-resolution T1-weighted structural image was obtained using a magnetization-prepared rapid acquisition gradient echo (MPRAGE) three-dimensional protocol (repetition time [TR] = 2400 ms, echo time [TE] = 2.22 ms, inversion time = 1000 ms, field of view [FOV] = 256 x 240 mm², flip angle = 8°, and 0.8-mm isotropic voxel). For functional imaging data, the participants underwent task-based fMRI (tb-fMRI) with multiband (anterior-posterior phase-encoding; acceleration factor = 8) interleaved acquisitions (T2-weighted EPI scans, TR = 800 ms, TE = 37 ms, 72 slices, slice thickness = 2 mm, FOV = 208 x 208 mm²). Tb-fMRI were acquired during three language and three motor paradigms. Production language (phonemic and semantic) tasks entailed 373 volumes, while the remaining tb-fMRI acquisitions entailed 223 volumes. Task procedures are described in the following section.

To assess language function, the tasks administered included: (1) Phonemic fluency: participants were instructed to generate as many words as possible starting with a specific letter (namely, “F”, “A”, “S”, “M”, and “E”, in this same order). (2) Semantic fluency: participants listed objects associated with specific environments (namely, school, kitchen, car, house, and hospital). (3) Auditory comprehension task: participants listened to a story narrated in a made-up language, and right after they listened to a story narrated in Spanish. Each of these tasks was repeated three times. To assess motor function a series of specific tasks targeting different movement patterns were implemented as follows: (1) Finger tapping task: participants tapped each finger against their thumb, one hand at a time, performing the task separately with the left and right hand. (2) Ankle flexion task: participants moved each foot up and down slowly, performing the task separately with the left and right foot. (3) Tongue movement task: participants moved their tongue in circular motions while keeping their mouth closed.

#### 2.3.2 fMRI preprocessing

Task-based fMRI was processed and analyzed using statistical parametric mapping (SPM12; Wellcome Trust Centre for Imaging Neuroscience, http://www.fil.ion.ucl.ac.uk/spm/) and MAGIC fMRI toolbox18, an in-house application developed with MATLAB 2017b. Prior to statistical analysis, functional scans from each subject underwent the standard fMRI preprocessing pipeline within the toolbox, involving the following steps: I) realignment of fMRI data using the mean image as a reference to remove any minor motion-related signal change and discarding any subject with a displacement greater than 3mm, II) slice-time correction, III) coregistration of the mean functional image to his corresponding structural image, IV) normalization into standard space (2×2×2mm; Montreal Neurology Institute, MNI coordinates) by using deformation fields obtained from the segmentation of the structural image, V) spatial smoothing with a Gaussian kernel of 8mm full width at half maximum (FWHM).

A first-level analysis was performed by fitting a general linear model on a voxelwise basis to model the brain activation of each task. Condition-specific effects were modeled by creating a boxcar function convolved with a canonical hemodynamic response function (HRF). Additionally, motion parameters were entered as regressors of no interest. Contrast images were calculated by subtracting the rest condition from the task condition. A criterion for statistically significant activation was set at a threshold of p-value < 0.001 and a family-wise error cluster threshold of p-value < 0.01 was applied to correct for multiple comparisons. All images were visually inspected for potential artifacts or technical problems. The size of significant clusters of activation reported by SPM were further used to evaluate NIP effect.

### 2.4 Subject classification and targeting procedures

Participants first underwent task-based fMRI to map functional activation (e.g., language function) allowing identification of critical functional regions at risk due to their proximity to the lesion. For each patient, we primarily used these fMRI activation maps to select NIBS targets. The primary approach was to place the stimulation site over peak fMRI activation clusters within the peritumoral region. When this approach was not possible (e.g., the main fMRI cluster was inaccessible to NIBS), we chose an alternative site within the same fMRI map, prioritizing both network relevance and peritumoral area proximity. In a subset of patients, the stimulation site was defined independently of fMRI: in cases 12, 15, and 17 it was localized using TMS motor mapping over M1, and in case 13 it was selected anatomically within the premotor/M1 region.

Following this assessment, participants were divided into two groups according to the targeted stimulation point (see Table 1). The language group (n=11) included individuals in whom language production networks were at risk (phonemic and semantic fluency); in these cases, language-related fMRI clusters were targeted with NIBS. The non-language group (n=15) comprised participants with motor areas at risk; here, stimulation was delivered to motor targets.

### 2.5 Non-invasive NIP intervention

The non-invasive NIP intervention program combined inhibitory-like neuromodulation targeting critical peritumoral brain activity with intensive task training, aiming to promote topographical reorganization of functional networks and facilitate the displacement of activity away from the lesion site. When rTMS was used, task training started immediately after the stimulation block. Targets were defined on each participant’s native structural MRI and stimulation was delivered using MRI-guided neuronavigation (Brainsight; Rogue Research, Montreal, Canada); details are provided in Boccuni et al. (2023). When tDCS was used, task training was performed concurrently during stimulation. Participants were scheduled to undergo between 10 and 20 treatment sessions. Sessions were provided either once or twice a day depending on the participant’s specific needs and schedule. Each session, lasting approximately one hour, was carefully personalized to suit the individual participant’s needs and abilities. Participants 12 and 13 completed a reduced number of treatment sessions, as their data were acquired during the initial pilot phase of the trial and are classified as part of a preliminary cohort comprising the first 10 participants enrolled in our prehabilitation program (Boccuni et al., 2024; see Table 1).

The primary tasks employed to train language functions comprised phonemic fluency, semantic fluency, categorization exercises, word games, completion of incomplete words, generation of novel words by altering letters in a given word, word list recall, and the narration of a film or story while alternating between languages (typically Catalan, Spanish, and English). Classic games, such as hangman and word search, were also incorporated. Furthermore, the method of loci was employed to enhance memory and language associations (Sousa et al, 2021; de Lange et al, 2017). The tasks were conducted using both conventional pen-and-paper techniques and the Guttmann NeuroPersonalTrainer® (GNPT; Solana et al., 2014), a validated online neurotraining platform. To promote progressive learning, the level of difficulty for each task was systematically increased both within each session and across the entire training program.

### 2.6 fMRI analysis

The FMRIB Software Library (FSL, version 5.0.10; https://fsl.fmrib.ox.ac.uk/fsl/fslwiki/) and Statistical Parametric Mapping (SPM, version 12; https://www.fil.ion.ucl.ac.uk/spm/) were used for analyzing MRI data.

The primary objective of this study was to assess NIBS-induced modulation of the language network, as reflected in language-related fMRI activity maps. To determine whether any observed effects were specific to stimulation of this network, we implemented complementary control analyses. In the language group, NIBS targeted the language network, and the motor network was included as a within-subject control to evaluate the anatomical and functional specificity of the effects. In the non-language group, NIBS targeted the motor network, providing a between-group control condition; in this group, language-network activity served as a reference to assess whether changes in the language group were attributable to direct stimulation rather than nonspecific or global effects. Across analyses, all motor-network measures were treated as control comparisons to strengthen inferences about the specificity of modulation within the language network. Analyses on these regards were conducted at two levels. First, a local effects analysis employed a region of interest (ROI)–based approach to evaluate neural changes at the stimulation target site pre- and post-NIP. Second, a network-level analysis compared the overall extent of task-related fMRI activation maps across individuals before and after NIP.

#### 2.6.1 Local effects of NIP at the stimulation target

To investigate local fMRI effects of the NIP intervention, we defined a spherical ROI (10-mm radius) centered on each individual stimulation target. A 10-mm sphere was chosen to balance anatomical specificity with robust sampling in normalized fMRI data: it provides sufficient voxel coverage for typical whole-brain resolutions (≈2–3 mm) while accommodating residual spatial uncertainty introduced by inter-individual anatomical variability, spatial normalization error and physiological/spatial dispersion of BOLD responses. Therefore, a 10-mm ROI was used to capture a reliable sample of activity in tissue immediately surrounding the stimulation site. In the language group, the ROI was centered on each participant’s stimulation target within their language-production network. The target was defined individually from pre-NIP task-based fMRI using two fluency paradigms (phonemic and semantic), and activation maps from both tasks were combined to delineate the participant’s language-production network The target was selected following the procedure detailed in Section 2.4.In the non-language group, the ROI was positioned over the individual stimulation target within the motor network, defined using pre-NIP task-based fMRI localizing motor activation. For within-subject specificity control analyses, an additional ROI was placed over the principal activation peak of the non-targeted network (i.e., the motor network in the language group and the language network in the non-language group) following the same functional localization and target-selection procedure.

#### 2.6.2 Effects of NIP on targeted network size

Changes in the spatial extent of functional network activity were assessed by comparing the number of activated voxels in the fMRI maps between pre- and post-NIP time-points. These analyses were conducted separately for both the language and motor networks in both the language and the non-language group.

To quantify these changes, we calculated the total volume of each network fMRI activation clusters (*V_net_*) by summing the number of suprathreshold voxels (*S_k_*) within the relevant fMRI activation clusters and multiplying by the voxel size, 2×2×2 mm³(*V_vox_*):

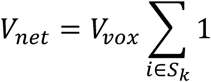

This volume served as an estimate of each participant’s individual language and motor network overall size and provided a basis for comparing network extent across NIP time points.

See Figure 2 for the representation of the fMRI analyses performed to assess the effects of NIP.

**Figure 2.**
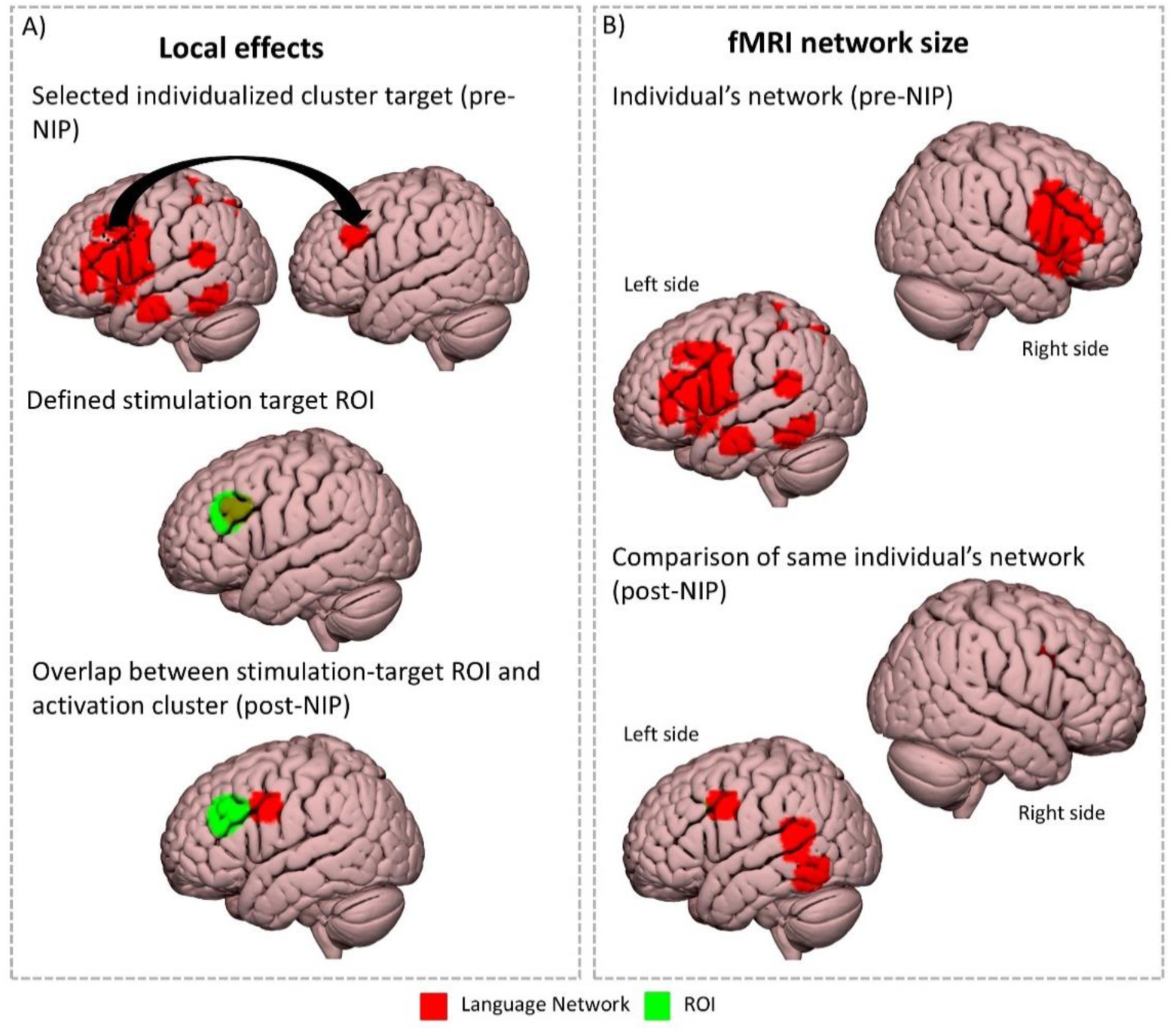
Schematic representation (illustrative example) of the main fMRI-based analyses performed to assess the effects of NIP. **A.** Local effects analysis: Individualized stimulation targets were identified within pre-NIP task-based fMRI activation clusters for language production (shown in red). A 10mm-radius spherical ROI (shown in green) was defined around this peak and later used to evaluate pre/post overlap of functional activation in the same region. **B.** Comparison of fMRI network size: Language-related activation maps from task-based fMRI (phonemic and semantic fluency tasks) were compared before and after the NIP intervention. The total number of suprathreshold voxels was computed for each individualized language network (shown in red), providing a quantitative measure of network extent.

### 2.7 Statistical analyses

Statistical analyses were conducted using R version 4.2.2 (R Core Team, 2022) via RStudio version 2022.07.1+554 (RStudio Team, 2022). For normally distributed data, parametric tests were used: t-tests (paired or independent, as appropriate) and mixed-design ANOVA for comparisons involving more than two groups or conditions. When the normality assumption was violated, nonparametric alternatives were applied: Wilcoxon signed-rank tests for paired comparisons and Wilcoxon rank-sum (Mann–Whitney U) tests for independent samples. Statistical significance was set at p ≤ 0.05.

#### 2.7.1 Neuroimaging-derived statistical analysis

##### 2.7.1.1 Local effects at the stimulation target

To investigate changes in brain activity at the stimulation site, we measured neuroimaging-derived data from the spatial overlap between the 10 mm-radius ROI (centered on the stimulation target) and the selected fMRI activation cluster in the targeted region at two time points (pre- and post-NIP). A mixed design ANOVA was conducted with group (language vs. non-language) as a between-subject factor, and time (pre-NIP vs. post-NIP) as within-subject factor, including both groups in the same model. Significant effects were further examined using post-hoc paired-samples t-tests (to assess within-group changes over time) and independent-samples t-tests (to compare groups at each time point).

##### 2.7.1.2 Changes in network size

To evaluate changes in the size of task-related networks following NIP, suprathreshold activation maps were generated from the SPM first-level contrasts using a voxel-wise family-wise error–corrected threshold of T > 4.69 (p < 0.01 FWE) and a minimum cluster extent of k ≥ 30 voxels. Thresholded, binarized maps were combined (summed) in FSL (v1.11.0) and then imported into MRIcroGL (v1.2.20220720) for volumetric quantification. For each participant, the volume (mm³) of the language and motor networks was computed as the number of suprathreshold voxels multiplied by voxel volume. A mixed design ANOVA was then performed from this neuroimaging-derived data with group (language vs. non-language) as a between-subject factor, and network (targeted vs. control) and time (pre-NIP vs. post-NIP) as within-subject factors.

##### 2.7.1.3 Descriptive analysis of the language network

To further characterize NIP-related changes in the language-network, which was the focus of this investigation, we conducted an additional qualitative, case-by-case inspection of fMRI activation patterns in the 11 participants assigned to the language group. This descriptive assessment allowed us to explore patterns beyond the overall amount of activation within the language network. Given the modest sample size and clinical heterogeneity, this qualitative approach was well suited to capturing patient-specific reorganization patterns. This analysis complements the quantitative results by describing spatial reorganization patterns that cannot be captured by volume-based statistics alone. All qualitative ratings were performed independently by two authors (N.B.-B.; A.R.-V.).

#### 2.7.2 Cognitive performance statistical analysis

To examine potential effects of NIP on cognitive performance, statistical analyses were conducted on both language and non-language tasks, as previously described in Section 2.2.1. Specifically, paired-sample t-tests were used to compare pre- and post-intervention scores for each cognitive measure. Language tasks were analyzed to monitor changes in the targeted domain, while non-language tasks (i.e visuospatial constructional and perceptual abilities) were included to assess possible transfer effects beyond language. Measures of central tendency and dispersion were reported using the median and interquartile range (IQR). Wilcoxon signed-rank tests were used for paired comparisons, and Kendall’s tau was used to assess associations between variables.

## 3. Results

### 3.1. Topographical changes in functional brain networks

#### 3.1.1 Local effects to the stimulation target

There was a significant interaction between group (language vs. non-language) and time (pre-vs. post-NIP) in ROI–activation cluster overlap within the language network, *F*(1,23) = 4.61, *p* = .04 (see Figure 3A for an illustrative example), indicating that the change over time differed between groups. Post-hoc analyses showed a significant reduction in overlap from pre- to post-NIP in the language group, *t*(10) = 5.68, *p* < .001, *d* = 1.71, whereas the non-language group showed no change, *t*(13) = 0.50, *p* = .62, *d* = 0.14 (Figure 3.B1). Between-group comparisons indicated no baseline difference, *t*(23) = 0.57, *p* = .57, but a significant group difference emerged post-NIP, *t*(23) = 2.43, *p* = .02. Analyses focused on the motor network revealed no effects of NIP on ROI–activation cluster overlap. In the non-language group, stimulation of the motor network did not induce changes in ROI–cluster overlap (no main effect of time: *F*(1,21) = 0.13, *p* = .72; no group × time interaction: *F*(1,21) = 0.62, *p* = .44). Similarly, showed no significant change in overlap within the hand-related motor cluster between pre- and post-NIP, *t*(9) = −0.60, *p* = .56, *d* = −0.19 (Figure 3.B2).

**Figure 3.**
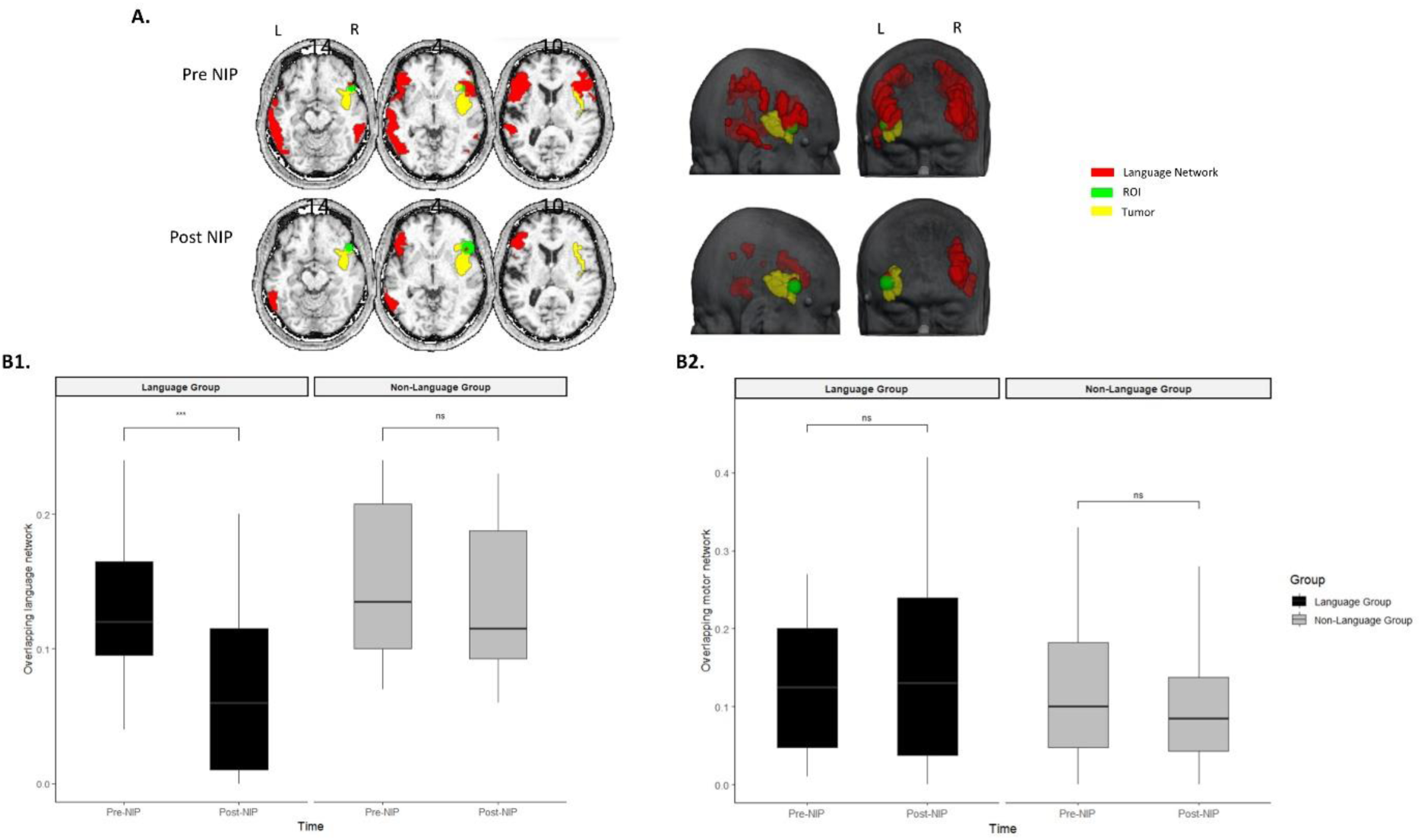
**A.** Example of the activation in fMRI language tasks: Pre vs Post intervention. In yellow the tumor, in red the task-related language network and in green the 10mm-radius ROI on the stimulation target. Shown here is Case 7 with a right-hemisphere tumor; note that task-related language activations (phonemic and semantic fluency tasks) can also be observed in the right hemisphere, adjacent to the tumor. **B1**. Comparison of the overlapping of language network within the ROI, for the language and non-language group. In both groups the ROI’s localization was within language network. Overall, a significant reduction of the overlapping and extent network size is shown in the language group, but not in the non-language group. **B2.** Comparison of the overlapping of motor network within the ROI, for the language and non-language group. In both groups the ROI’s localization was within motor network. The asterisk indicates a significant main effect of Time (pre- vs post-NIP) on network size across groups. Asterisks indicate statistical significance levels: * p < 0.05, ** p < 0.01, *** p < 0.001; ns = not significant.

#### 3.1.2 Comparison of overall fMRI network size

For the language network, there was no significant interaction between group (language vs. non-language) and time (pre- vs. post-NIP) on task-related language network size, *F*(1, 24) = 1.02, *p* = .32, indicating that volume changes over time did not differ between groups. However, there was a main effect of time, with network size decreasing from pre- to post-NIP across participants, *F*(1, 24) = 5.34, *p* = .03, consistent with a general reduction in the extent of task-related activation over time. Additional illustrations of the activation of language production clusters before and after prehabilitation in the language group (language network) and the non-language group (language network) are provided in the Supplementary Material (Figures S1–S2). Analyses of the motor network revealed no significant effects on motor network size (main effect of time: *F*(1, 23) = 0.74, *p* = .40; group × time interaction: *F*(1, 23) = 0.03, *p* = .87; main effect of group: *F*(1, 23) = 0.54, *p* = .47), indicating stable motor-network size across time and groups.

#### 3.1.3 Descriptive analysis of overall fMRI language network size

Reorganization patterns were heterogeneous (Table 2). Some participants showed intra- and/or interhemispheric recruitment, whereas others showed localized intrahemispheric shifts or no clear cross-hemispheric reorganization. Activation clusters shifted anteriorly, posteriorly, or in mixed directions, generally moving away from the tumor. Intrahemispheric network volume decreased in most participants (9/11), while interhemispheric volume changes were more variable (see Supplementary Material, fMRI language network results).

**Table 2.**
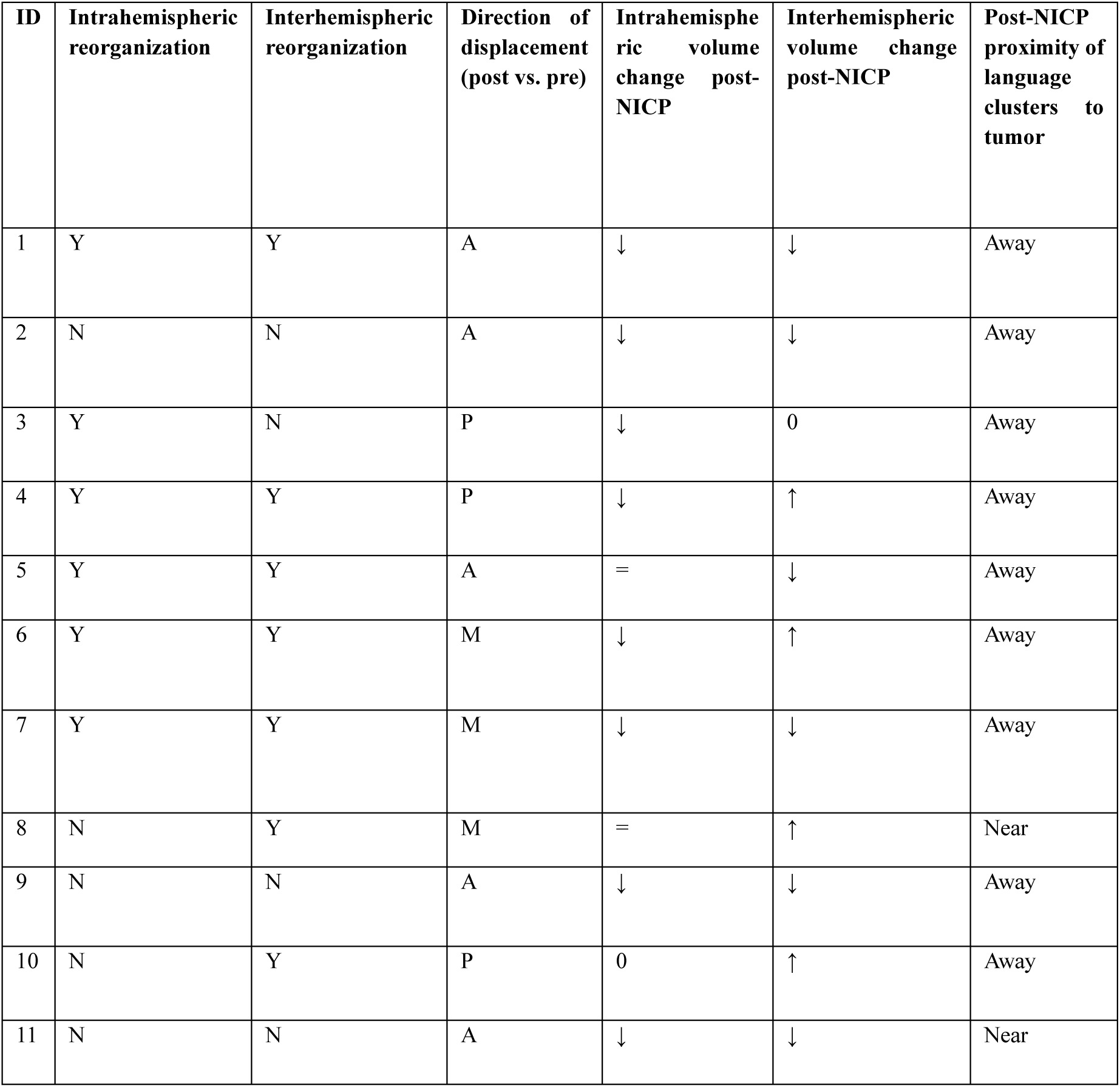
Quantitative description of the language network in the 11 cases of the language group. For each participant, the table reports intrahemispheric and interhemispheric reorganization, direction of displacement, changes in intra- and interhemispheric network volume, and the post-NICP proximity of language-related clusters to the tumor. Coding scheme: intrahemispheric/interhemispheric reorganization – Y = present, N = absent; direction of displacement – A = predominantly anterior, P = predominantly posterior, M = mixed anterior–posterior; volume change (intra-/interhemispheric) – ↓ = decreased post-NICP, ↑ = increased post-NICP, = = no relevant change, 0 = no suprathreshold activation; proximity to tumor (post-NICP) – Near = clusters remain close to the peritumoral region, Away = clusters clearly displaced from the peritumoral region.

### 3.2 Language and other cognitive domain performance results

Clinical outcomes are presented in Table 3. Language performance remained stable in both the language and non-language groups, with no significant changes observed (all p’s > 0.05). Furthermore, non-language domains, including visuospatial constructional and perceptual abilities, also showed no significant differences in all groups before and after the NIP intervention (all p’s > 0.05).

**Table 3.** Overall cognitive outcomes pre-NIP and post-NIP. Values are presented as p-value, mean (standard deviation). NA: p-value could not be computed due to absence of variability (identical scores pre- and post-NIP).

| whole cohort (n = 26) |  |  |  |  |
| --- | --- | --- | --- | --- |
|  | language group (n = 11) |  | non-language group (n = 15) |  |
| Clinical assessment, cognitive function | p-value* | Mean (SD) | p-value* | Mean (SD) |
| TB-R (conversation and narration) | NA | 7.64(1.2) | 1.00 | 8(0) |
| TB-R (thematic narration) | 1.00 | 5.73(0.9) | 1.00 | 5.87(0.35) |
| TB-R (picture description) | 0.37 | 5.64(0.92) | 1.00 | 5.73(0.59) |
| TB-R (verbal word repetition) | 1.00 | 9.36(2.11) | 1.00 | 9.93(0.26) |
| TB-R (verbal naming) | 1.00 | 5.73(0.9) | 1.00 | 6(0) |
| TB-R (verbal comprehension) | 1.00 | 15.27(1.85) | 1.00 | 16(0) |
| TB-R (reading comprehension) | 1.00 | 7.82(0.6) | 1.00 | 8(0) |
| TB-R (automatic language) | 0.82 | 2.73(0.90) | 0.06 | 2.78(0.8) |
| Forward | 1.00 | 2.6(0.97) | 0.61 | 2.75(0.82) |
| Reverse |  |  |  |  |
| TB-R (verbal repetition) | NA | 8(0) | 1.00 | 8(0) |
| Syllables | 1.00 | 7.36(2.11) | 1.00 | 8(0) |
| Logatomes | 1.00 | 55(16.58) | 1.00 | 60(0) |
| Sentences |  |  |  |  |
| TB-R (naming) |  |  |  |  |
| Visuoverbal naming | 0.59 | 13.81(0.41) | 1.00 | 13.73(1) |
| Naming completion | 1.00 | 5.72(0.9) | 1.00 | 6(0) |
| TB-R (visuospatial attention) | 1.00 | 28(0) | 0.59 | 27.93 (0.27) |
| WAIS IV blocks (visuoconstruction) | 0.34 | 32.86(10.38) | 0.33 | 38.77(9.6) |

*\*p-values were obtained from Wilcoxon signed-rank (paired) tests comparing pre-NIP vs post-NIP scores within each group. Abbreviation: TB-R, Barcelona-Revised Test.*

## 4. Discussion

This study shows that preoperative NIP combining non-invasive neuromodulation (rTMS and/or tDCS) with intensive language training can modulate the language network in patients with brain tumors. Specifically, we observed a reduction in peritumoral language-related activation without comparable effects in the motor network, indicating network-specific reorganization. Importantly, this neuromodulation occurred without deterioration in language or broader cognitive performance, supporting the capacity of targeted NIP to guide selective neuroplasticity prior to surgery.

### 4.1 Specific reorganization of the language network through NIP

Our neuroimaging results revealed selective modulation of the language network in the language-targeted cohort, supporting the hypothesis that network-targeted NIP procedures can drive topographically specific reorganization within multimodal functional networks, with significant translational relevance in brain tumor surgery. In this study, we observed a consistent reduction in the spatial extent of fMRI activation clusters within the NIBS target region, which was defined based on individual critical fMRI language maps, supporting successful topographical reorganization of the language network. Indeed, the combination of local fMRI signal reduction and preserved performance is consistent with adaptive neuroplasticity mechanisms described in prior work, where stimulation of critical areas is associated with compensatory recruitment of alternative, structurally preserved regions (Hartwigsen, 2021; Nieberlein et al., 2023).

Notably, the imaging changes following NIP were specific to the targeted language network, as confirmed by control analyses (Momi et al., 2021a; 2021b). Because stimulation was combined with language training that engages the same network, the relative contribution of each component cannot be disentangled. Nevertheless, the observed reduction in peritumoral language activation and the broader topographical reorganization within the network are consistent with the goals of prehabilitation: decreasing reliance on vulnerable cortex adjacent to the tumor while preserving function (Boccuni et al., 2023).

### 4.2 Reorganization patterns in fMRI activation maps

The ROI analysis identified a significant reduction in peritumoral language activation following NIP, representing the primary effect of the intervention. Descriptive analyses further suggested redistribution of activation away from peritumoral regions, with heterogeneous intra- and interhemispheric patterns across participants. Although exploratory, these changes are consistent with mechanisms of remapping, functional unmasking (Pascual-Leone et al., 2005), and compensatory recruitment reported in brain tumor populations (Duffau, 2005). Together, these findings extend prior evidence of tumor-related network reorganization (Aerts et al., 2016; Morell et al., 2022; Manso-Ortega et al., 2023) by indicating that neuromodulation combined with training may further modulate language network topography.

### 4.3 Multimodal vs. unimodal network responsiveness to NIP

In the present study, we observed that multimodal or higher-order networks, such as the language network, are differentially modulated compared with unimodal or low-order sensorimotor systems. A key theoretical implication is that multimodal association networks may be particularly amenable to targeted neuromodulation and topographical reorganization, likely due to their distributed cortical architecture. The language network integrates information across multiple cortical regions, whereas primary motor networks are more spatially constrained and somatotopically organized (Yeo et al., 2011; Mesulam, 1998; Duffau, 2014). Recent evidence supports this distinction: hierarchical neurodevelopment prolongs plasticity windows in association cortices, enabling sustained flexibility in multimodal regions (Sydnor et al., 2021), and neocortical gradients reveal tighter structure–function coupling in unimodal areas versus more flexible alignment in transmodal regions, facilitating reconfiguration after perturbation (Vázquez-Rodríguez et al., 2019). Stimulation mapping further demonstrates greater excitability and inter-network integration in higher-order systems (e.g., language, default mode) compared to sensorimotor networks (Momi et al., 2025), with rapid short-term reorganization observed following neuromodulation (Hartwigsen et al., 2017). Consistent with these frameworks, language-targeted NIP in our study produced measurable network-specific reorganization, whereas motor-targeted NIP (which can also entail potential neural and clinical effects) did not produce similar modulations measurable with a 10 mm ROI and whole-network size. This pattern observed in our data aligns with models of high-order plasticity mechanisms and is consistent with the notion that multimodal systems exhibit higher network plasticity and greater flexibility for reconfiguration following perturbation, while unimodal systems show greater functional stability and constraint (Gatica et al., 2025; Cramer et al., 2011).

### 4.4 Targeted neuromodulation and language network plasticity

Selective reorganization of the language network following NIP is consistent with models of experience-dependent plasticity, in which repeated modulation combined with task engagement promotes redistribution within large-scale networks. The multi-session, individually targeted stimulation approach used here aligns with prior evidence showing that repeated neuromodulation can reinforce plastic changes and enhance network reorganization over time (Klooster et al., 2023; Maeda et al., 2000; Wang et al., 2014; Liu et al., 2018; Lefaucheur et al., 2020). Given the known variability in neuromodulation outcomes (López-Alonso et al., 2014; Hamada & Rothwell, 2015; Perellón-Alfonso et al., 2018), personalized targeting and repeated sessions appear particularly important for achieving reliable network-level effects. These approaches may help stabilize network reorganization and ensure meaningful clinical benefits (Maeda et al., 2000; Klooster et al., 2023). Notably, in our study modulation was observed despite tumor–brain crosstalk and anatomical constraints (Ghinda, Wu & Duffau, 2023), suggesting that TMS and tDCS can influence functional organization even in structurally compromised systems (Boccuni et al., 2024a; 2024b). Our findings suggest that NIP may facilitate reorganization process and presumably support a more favorable network configuration ahead of surgery.

### 4.5 Language network modulation and cognitive preservation across NIP intervention

Neuroimaging effects were confined to the targeted language network, with no evidence of modulation in non-targeted systems. Both between-subject comparisons and within-subject controls confirmed this specificity. These observations align with prior NIBS studies demonstrating that modulation of specific neural nodes on concrete brain networks produces specific effects within these brain networks, which can be captured with functional neuroimaging techniques (i.e., Abellaneda-Pérez et al., 2019; Momi et al., 2021b; Wang et al., 2014). The use of targeted, functionally unrelated regions (i.e. motor) as control conditions aligns with strategies previously described in the literature, as those outlined by Hartwigsen (2016) on the possibility of stimulating neighboring or contralateral regions that are not expected to contribute to the task under investigation, to control for unspecific effects while preserving anatomical and procedural comparability across groups.

Interestingly, our observations indicated a general reduction in the overall size of task-related activation across both groups. Because participants completed identical fMRI tasks at two time points, this pattern is plausibly consistent with practice-related effects and repetition suppression (Gobel et al., 2011; Ewbank et al., 2011). The phenomenon of repetition suppression refers to reduced neuronal activation upon repeated exposure to the same stimulus, indicating a refined engagement of neural resources, rather than a loss of function (Stam et al., 2021; Lee, Henson, & Lin, 2020; Barron et al., 2016; Vannini et al., 2012). In this context, reduced activation may reflect a more economical allocation of neural resources during repeated task performance rather than diminished functional engagement.

Moreover, and in association with the previous neural findings, language performance remained stable across NIP intervention, suggesting that the induced reorganization at the brain level did not compromise behavioral outcomes. The implications of reduced task-related activation observed in our participants highlight a trend where fewer resources may be utilized for enhanced processing efficacy, echoing findings in prior research demonstrating that participants with greater task familiarity or procedural learning may exhibit diminished activation within targeted neural circuits (Larsson & Smith, 2011; Epstein et al., 2008). The stability of cognitive performance supports this interpretation, suggesting that smaller activation patterns may reflect refined, rather than diminished, functional engagement.

### 4.6 Network plasticity in a preoperative context

These findings indicate that network plasticity remains accessible in brain tumor patients and can be modulated through targeted neuromodulation combined with training. The integration of NIBS and neuroimaging may therefore provide a framework for guiding adaptive reorganization prior to surgery (Zhang et al., 2021; Pi et al., 2019; Voss et al., 2010). Preservation of language and non-language performance suggests that such modulation can occur without behavioral compromise, a critical consideration in brain tumor patients vulnerable to crowding effects or maladaptive reorganization (François et al., 2019; Martínez-Lozada et al., 2025).

## 5. Limitations

The first limitation of this study was that the data comes from a phase 1 clinical trial and from a previous pilot phase (Boccuni et al., 2023), conducted as a case series without a control group. To mitigate this limitation, we implemented internal control strategies: non-targeted networks served as controls to verify that the observed effects were specific to the targeted system. Additionally, the sample size was small, particularly after stratification into language and non-language groups. Recruitment for preoperative neuromodulation protocols in brain tumor patients is inherently challenging due to clinical and logistical constraints. In addition, the cohort was clinically heterogeneous, reflecting variability in tumor location, tumor characteristics, and baseline patient status. Another important limitation concerns the inability to disentangle the relative contributions of neuromodulation and intensive language training. Because stimulation was systematically paired with training, their independent effects cannot be isolated. It is plausible that the local reduction of activation near the stimulation target was primarily driven by neuromodulation, whereas the broader topographical redistribution observed in descriptive analyses may reflect training-related influences. With respect to language performance, it is important to note that language assessment was not performed during brain activity acquisition via MRI; instead, it was performed offline using pen-and-paper evaluations in the days surrounding the MRI sessions. The observed ceiling effects in language-cognitive assessments are more plausibly attributable to the limited sensitivity and variability of the tests employed, highlighting the need for future studies to incorporate more refined language evaluation tools.

## 6. Conclusions

In conclusion, our findings suggest that preoperative NIP can selectively reorganize language network in brain tumor patients while preserving cognitive performance. The absence of modulation in non-targeted networks supports the specificity of this effect. This targeted modulation highlights the potential to guide adaptive neuroplasticity prior to surgery, potentially reducing functional constraints and supporting safer, more extensive resections in language-critical regions.

## Acknowledgments

The authors express their gratitude to all participants of the Prehabilita program, as well as to the dedicated research staff and neurosurgical units involved in this study. We are also indebted to the Magnetic Resonance Imaging Core Facility of the Institut d’Investigacions Biomèdiques August Pi i Sunyer (IDIBAPS) for the scientific and technical support in MRI acquisition and analysis.

## Data availability

To protect participant confidentiality and in accordance with ethical approval and informed consent, the datasets from this study are not publicly accessible. Access is restricted and may be granted upon reasonable request. For further details, please contact the corresponding authors.

## Ethics approval and consent to participate

All procedures from the present study were performed in accordance with the Helsinki declaration. The study was approved by the Research Ethical Committee of Fundació Unió Catalana d’Hospitals (approval number: CEI 21/65, version 1, 13/07/2021). Clinician participants provided informed consent by reviewing and agreeing to a consent statement presented by the research team, prior to responding to any questions or proceeding with any treatment.

## Consent for publication

Participants in the study provided consent for the use of anonymized quotations in research publications. All identifying details have been excluded to ensure participant confidentiality.

## Author’s contributions

N.B.B. and A.R.V. contributed to investigation, formal analysis, data curation, visualization, and writing – original draft, writing – review & editing. J.M.T., K.A.P., and A.P.L. contributed to conceptualization, methodology, investigation, resources, and writing – review & editing. S.D.G. contributed to investigation, data curation, and writing – review & editing. K.A.P. contributed to project administration and supervision. E.B.O., R.P.A., and D.B.F. contributed to investigation and writing – review & editing. L.B. contributed to investigation. C.L., E.M.M., and N.B. contributed to software, methodology, and investigation. All authors read and approved the final manuscript.

## Declaration of generative AI and AI-assisted technologies in the writing process

During the preparation of this work the authors used [OpenAI. GPT-5.2 model] to support language editing and improve the clarity, readability, and structure of the manuscript text.

After using this tool/service, the authors reviewed and edited the content as needed and take full responsibility for the content of the publication.

## List of abbreviations

NIP: Neuromodulation-induced prehabilitation
TMS: Transcranial magnetic stimulation
tDCS: Transcranial direct current stimulation
tb-fMRI: task-based functional MRI
ROI: Region of Interest
CNS: Central nervous system
IFG: Inferior frontal gyri
pSTG: Posterior superior temporal gyri
NIBS: Non-invasive brain stimulation
iTBS: Intermittent theta-burst stimulation
TB-R: Barcelona-Revised Test

