## Supplementary Material for "Non-invasive prehabilitation before neurosurgery modifies the topography of brain language networks without compromising function"

### Representation of cluster activation

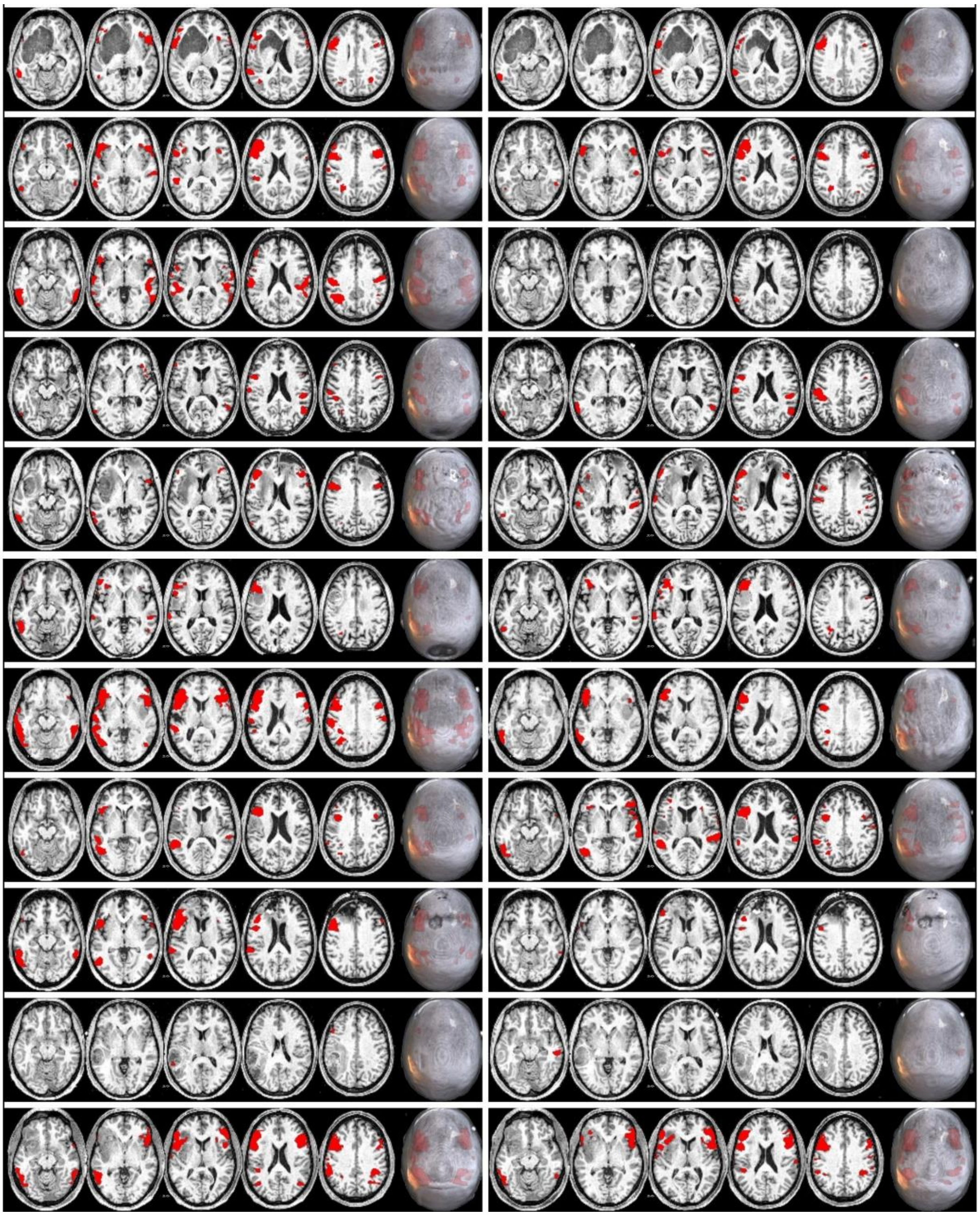

**Figure S1.** Activity maps of language production clusters before (left) and after (right) prehabilitation in the language group.

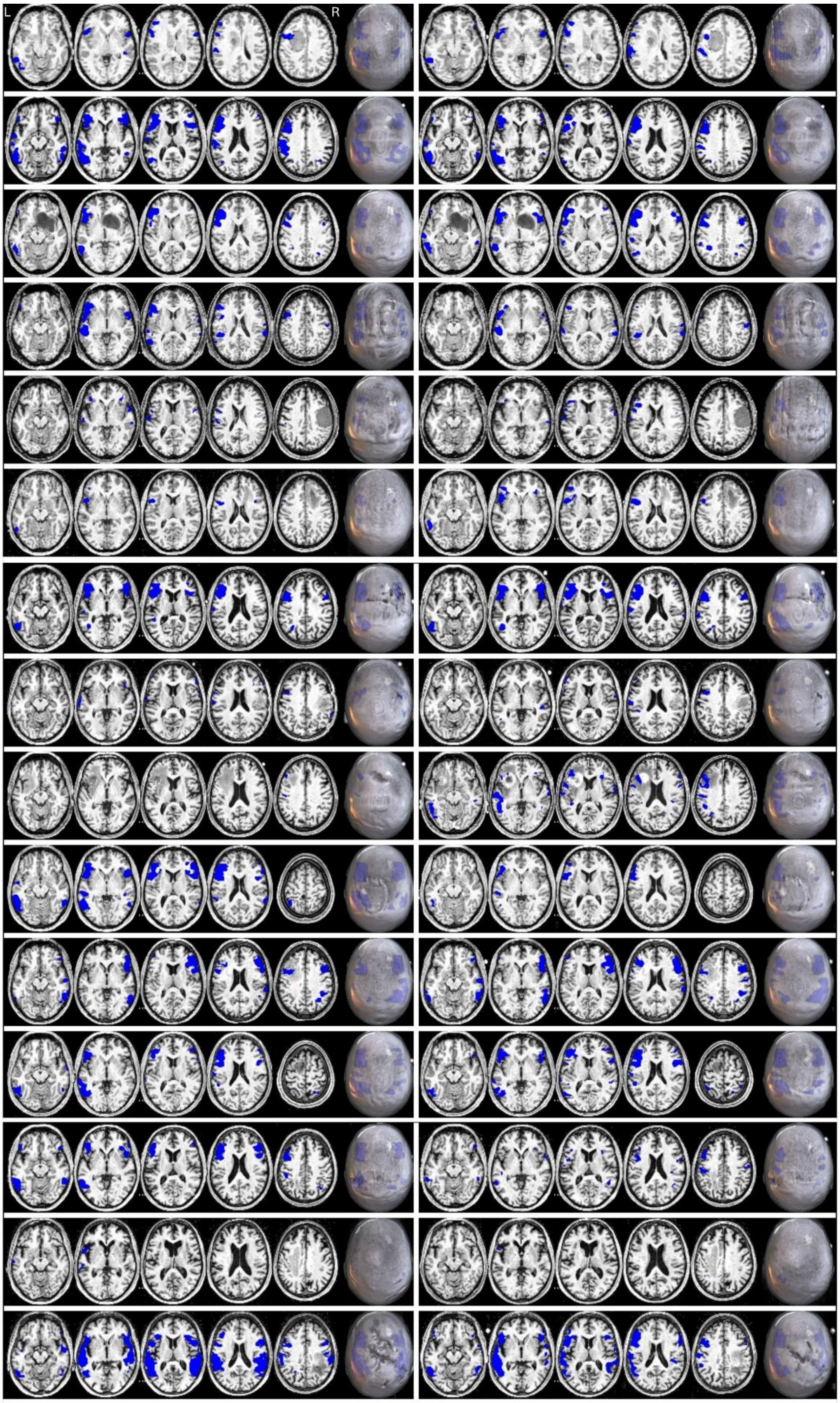


**Figure S2.** Activity maps of language production clusters before (left) and after (right) prehabilitation in the “non-language” group (control condition).

**fMRI language network descriptive results**

Regarding spatial reorganization, 1 of 11 participants showed predominantly interhemispheric recruitment, 1 of 11 showed intrahemispheric shifts, and 5 participants exhibited both types of reorganization. The remaining 3 participants did not show clear hemispheric-level reorganization. In terms of regional displacement, activity patterns moved in anterior or posterior directions depending on individual anatomy; however, a majority showed shifts away from the tumor or stimulation target. More specifically, 5 participants demonstrated predominantly anterior displacement, 3 showed posterior displacement, and 3 exhibited mixed anterior–posterior changes.

Changes in network volume were more heterogeneous. Interhemispheric volume decreased in 6 participants, increased in 4, and was absent in 1 at post-treatment. Intrahemispheric volume decreased in 9 of 11 participants, remained stable in 2, and was absent in 1. Only two participants showed post-treatment activation clusters that remained relatively close to the tumor, whereas in the remaining cases the language-related activity shifted clearly away from the peritumoral zone (see Table S1).
